# Psychometric characteristics of ADHD-T-5 in its Dari version among public primary school students in Kabul City, Afghanistan

**DOI:** 10.1101/2024.07.09.24309985

**Authors:** Ziauddin Azimi, Seok Mui Wang, Jesmine Khan, Zaliha Ismail, Mohamad Rodi Isa

**Affiliations:** Department of Biochemistry and Molecular Medicine, Faculty of Medicine, Universiti Teknologi MARA, Sungai Buloh Campus, Selangor, Malaysia; Department of Public Health Medicine, Faculty of Medicine, Universiti Teknologi MARA, Sungai Buloh Campus, Selangor, Malaysia; Department of Medical Microbiology and Parasitology, Faculty of Medicine, Universiti Teknologi MARA, Sungai Buloh Campus, Selangor, Malaysia; Institute of Pathology, Laboratory and Forensic Medicine (I-PPerForM), Universiti Teknologi MARA, Sungai Buloh Campus, Selangor, Malaysia; Non-Destructive Biomedical and Pharmaceutical Research Centre, Smart Manufacturing Research Institute (SMRI), Universiti Teknologi MARA, Puncak Alam Campus, Selangor, Malaysia; Department of Biochemistry, Faculty of Pharmacy, Kabul University, Jamal Mina, Kabul, Afghanistan

## Abstract

**Introduction:** One of the key instruments used to assess attention-deficit/hyperactivity disorder (ADHD) in children and adolescents is the ADHD Rating Scale-5. This study aimed to test the psychometric characteristics of the ADHD Rating Scale-5 teacher version (ADHD-T) in Dari among school-aged children in Kabul City.

**Methods:** Face and content validity, explanatory factor analysis (EFA), confirmatory factor analysis (CFA) and reliability of the questionnaire were conducted. The structure, validity, and reliability of the scales were evaluated using the translated versions of the ADHD-T questionnaires for the assessment of ADHD in children. Teachers completed the ADHD-T-Dari on behalf of three hundred and fifty-eight public primary school students.

**Results:** Face validity, content quality, and internal consistency of the ADHD-T-Dari were excellent. Significant collinearity was found between the items of the inattention and/or hyperactivity-impulsivity domains of the ADHD-T-Dari. The Kaiser-Meyer-Olkin index was 0.913 and was satisfactory. The two-factor model of the scale showed a better fit with an RMSEA of 0.065, a CMIN/*df* of 20.501, and a CFI of 905 compared to the one- and three-factor models of ADHD-T-Dari. The Cronbach’s alpha for the total ADHD-T-Dari scale with 18 items was 0.898, while it was 0.851 and 0.847 for the inattention and/or hyperactivity-impulsivity subscales with 9 items each.

**Conclusion:** The exploratory and CFA showed that the psychometric qualities have strong concept validity and very good psychometric properties with good reliability. Therefore, the ADHD-T-Dari is valid, reliable, and suitable for the assessment of ADHD symptoms in Afghan school children and adolescents.

## Introduction

Attention-deficit/hyperactivity disorder (ADHD) is a neurodevelopmental disorder characterised by persistent symptoms of inattention and/or hyperactivity-impulsivity that do not match the patient’s neurological stage. Approximately 2 to 7% of children and adolescents suffer from ADHD, making it one of the most common disorders in these age groups [1,2]. ADHD is primarily characterised by inattention, hyperactivity, or impulsivity. Therefore, a thorough assessment of symptoms should include a clinical interview with the patient and their parents, as well as the collection of data on the impact of symptoms on the patient’s overall performance, both at home and at school [3]. Although both hereditary and environmental factors are considered in the diagnosis, the exact origin of ADHD is still unclear. In addition to neuropsychiatric and biological changes, social factors may also have an impact on how severe the difficulties of the disorder are [4]. Children with ADHD have great difficulty controlling their behaviour in response to environmental stresses. These challenges typically lead to problems with behaviour regulation, academic performance, and interactions with peers and family members [5]. The Diagnostic and Statistical Manual of Mental Disorders, Fifth Edition (DSM-5), states that ADHD is characterised by two main symptoms, namely inattention and hyperactivity/impulsivity [6].

A planned diagnostic interview with the child, parents, and teachers, completion of ADHD rating scales by parents and teachers, and close observation of behaviour at school and during clinical testing are all components of an evidence-based assessment of ADHD [7]. Teachers have access to pertinent information about a student’s behaviour at school, including details about how they interact with classmates and other students [8]. Clinicians need to use reliable and validated instruments when assessing children and adolescents suspected of having ADHD, taking into account the frequency, severity, and wide range of socio-emotional and cognitive challenges associated with ADHD. Direct observation of behaviour at school and clinical testing are as much a part of an evidence-based assessment of ADHD as organised diagnostic interviews with the child, parents, and teachers [7]. The teacher’s assessment of the child’s behaviour is crucial. ADHD rating scales completed by parents and teachers are inexpensive, easy-to-use tools for diagnosing ADHD. Teachers have access to vital information about a student’s activities at school, including details about how he or she interacts with peers and other students. Research has shown that a teacher’s assessment is more reliable than a parent’s assessment while being more sensitive to hyperactive behaviour [9]. The ADHD Rating Scale-5 is an instrument that, due to its simple structure and method of administration, is well suited for intervention studies in which repeated administrations are used to assess changes in behaviour. Following the established DSM-5 criteria, ADHD-5 offers clinicians the opportunity to collect information from both parents (ADHD-5 Home version) and teachers (ADHD-5 School version) on the frequency of each of the defining symptoms of ADHD [7].

There are no validated screening instruments in the Dari language that could help teachers in Afghanistan to identify students with ADHD symptoms who may need further screening for learning problems. Although several teacher-completed questionnaires have been developed to assess ADHD symptoms and symptoms of other related mental health conditions, the main challenge in evidence-based assessment, according to international standards for the translation/adaptation of psychological testing instruments, is to validate the adapted scale in the culture in which it was originally developed. When using a scale, whether in research or clinical practice, it is crucial to demonstrate internal consistency. This is defined as the psychometric quality of a test that demonstrates equivalence in the latent variable under investigation [10,11]. The aim of this study was to validate the Dari-language vision of ADHD-T and to describe the prevalence of ADHD symptoms in public primary school children in Kabul City, Afghanistan.

## Materials and methods

### Sampling method

Three hundred and fifty-eight public primary school students from Kabul City, Afghanistan (174 males and 184 females), randomly selected from five schools and three districts, served as the study’s subjects. A printed version of the ADHD-5-Dari was used by 40 female teachers who had been teaching the students for at least six months to assess the students. Fig 1 shows how the study participants were included and excluded from the study. The study was conducted from March 2022 to August 2022. The students participated in the study were between 6 and 9 years old, with a mean (*M*) age of 7.7 years, and a standard deviation (*SD*) of 0.93. A total of 511 students (260 males and 251 females) were selected from 5 schools using simple random sampling. Due to disputes between parents, disagreements between children, and incomplete demographic information, 153 students (86 males and 67 females) were excluded.

**Fig 1.**
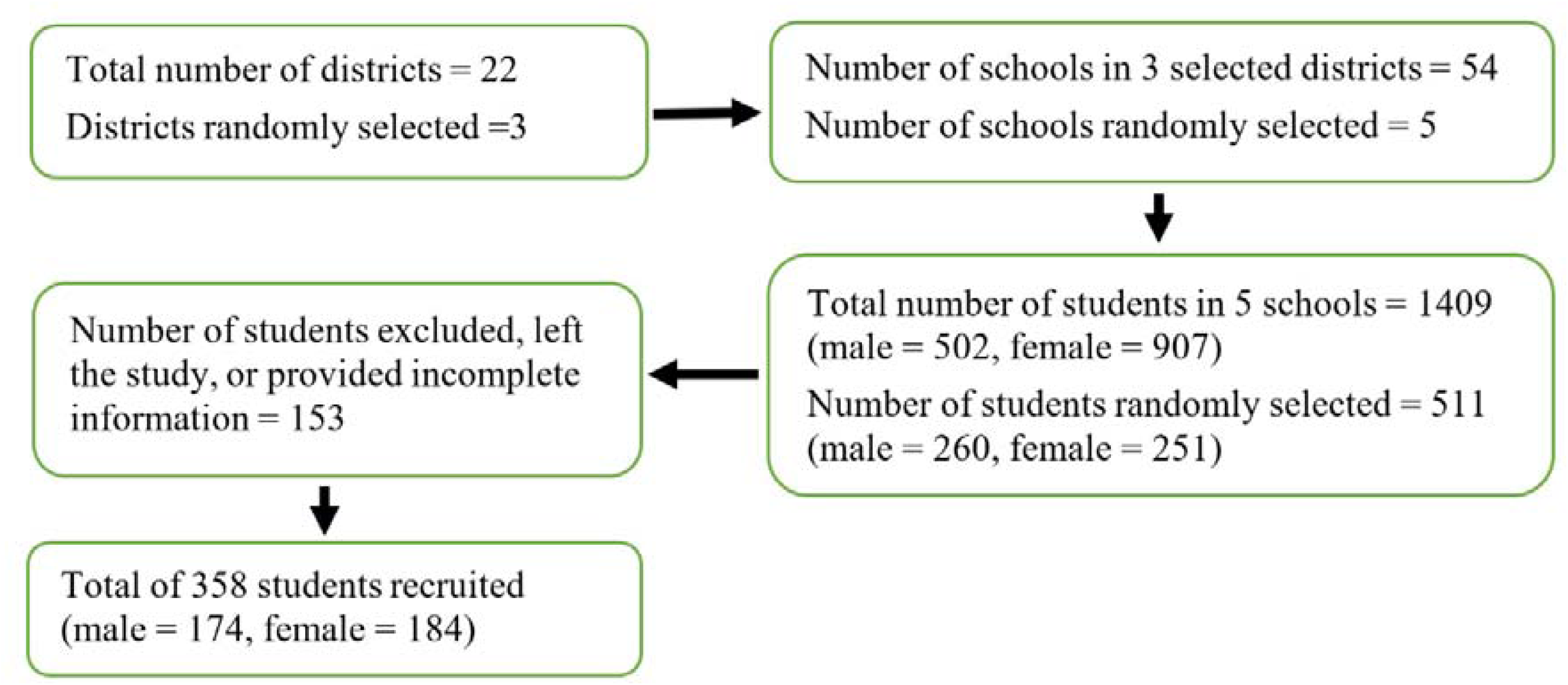
Sampling criteria for the selection of districts, schools, and students enrolled in this study.

### ADHD-T

The ADHD-T Rating Scale-5 was developed and updated by DuPaul *et al*. (2016). It is an 18-item scale for assessing children that reflects the symptoms of ADHD defined in the DSM-5. The original questionnaire has very good psychometric properties [7]. For the adaptation and validation of this questionnaire, written permission was obtained from the developer of the questionnaire by email. The symptom scale contains two subscales: one for inattention and one for hyperactivity-impulsivity. Each response is rated on a four-point Likert scale, where 0 is “never or seldom”, 1 is “occasionally”, 2 is “frequently”, and 3 is “very often”. The sums of all associated items are used to determine the total score for the (sub)scales. Teachers can use this scale to assess a student’s ADHD behaviour at school. This questionnaire has a high level of content, construction, and reliability (internal consistency). The validation of the 18 items of ADHD-T resulted in a two-dimensional validation. The reliability and validity of the instrument were consistent in children aged 5 to 17 years [7].

### Translation of questionnaire

In accordance with the standards established by Sousa & Rojjanasrirat [11], linguistic and cultural adaptations were made, and the psychometric characteristics of the ADHD-T Dari language versions were determined. The translation and validation process comprised four stages.

In the first stage, the ADHD-T [7] was translated into English by two different translators. The first translator was proficient in both the target language and the source language of the questionnaire and was multilingual with Dari as his native language. The translator had a good understanding of the colloquial language and everyday use of the target language but was not familiar with the terminology used in the questionnaire. The second translator was also multilingual and a native English speaker. He spoke both languages well and was also bilingual, but unlike the other translator, he was familiar with the terms used in the questionnaire. The translators’ efforts resulted in the production of two ADHD-T versions in Dari. In the second stage, the two translations served as the basis for the creation of one version of the questionnaire. The questionnaire was created with the help of the study team, the two translators, and a third translator. In the third stage, two independent translators with the same characteristics as those in stage 1, back-translated the version of the accepted questionnaire. None of the translators were familiar with the original ADHD-T text. As a result of the translators’ work, two back translations into English were produced. In the fourth stage, a multidisciplinary panel of experts reviewed all five versions of the questionnaires to determine the transcultural equivalence between the original ADHD-T and ADHD-T-Dari. The expert panel consisted of the members of the research team and five translators who had worked on the previous stages of the process. The members of the expert panel agreed and produced the final ADHD-T-Dari version, which was available for validation.

### Content and face validity

The item-level content validity index (I-CVI) and the scale-level content validity index (S-CVI) for the ADHD-T-Dari were calculated. Eight professionals, one with a PhD and seven with a master’s degree (four from the Department of Counselling and four from the Department of Psychology) from the Faculty of Psychological Science, Kabul University were asked to rate each item on a four-point Likert scale (1 = not important, 2 = somewhat relevant, 3 = fairly significant, and 4 = highly relevant) in the context of its relevance to understanding ADHD.

For the face validation of the ADHD-T-Dari questionnaire, 40 highly qualified teachers with at least a bachelor’s degree and 10 years of experience were chosen. The paper-based version of the questionnaire was distributed to the teachers. The questionnaire was self-administered, and participants were asked to note the time taken to complete it, the readability of the content, the language and terminology used, and the overall structure of the questionnaire. Their perceptions of how well they understood the questions and their content were rated and noted. This also included their comprehension of the language and overall design. Based on their comments and recommendations, minor corrections and fine-tuning were made to the questionnaire. It was then decided that a comprehensibility level of around 80% was considered acceptable [11].

### Data analysis

Descriptive statistics were used to assess student demographic background and practise data. Data were tested for normal distribution. Content validity (content validity index), face validity (face validity index), item analysis (discrimination index), internal consistency (Cronbach’s alpha), and theoretical relevance (exploratory and confirmative factor analysis) of the ADHD-T-Dari were assessed to evaluate psychometric characteristics.

For content validity, an I-CVI of 0.78 or more and an S-CVA/Ave of 0.90 or more were considered indicative of content validity [11]. Standards were then rated as fair, good, or excellent for each k value. The S-CVI/averages were determined by dividing the sum of the I-CVI by the total number of items. The I-CVI of each item was calculated by dividing the number of agreeing experts by the total number of experts. The probability of chance occurrence (Pc) was estimated as Pc = [N! / A! (N-A)!] × 0.5N, and the kappa was calculated as k = (I-CVI - Pc) / (1 −1Pc).

For face validity, interobserver agreement was then measured using the kappa index. Values between 0.61 and 0.80 were considered to indicate a respectable but acceptable level of agreement, while values of 0.81 or more indicated an above average level of acceptable agreement [11]. Finally, the item-level face validity index (I-FVI) and the scale-level face validity index (S-FVI) were calculated using a formula similar to the S-CVI and I-CVI.

Item analysis was used to separate the values of the calculated indices (inter-item correlations) for each item. Negative values were not allowed for the indices. The data collected was also checked to see whether any floor or ceiling impacts had occurred. An investigation of the internal consistency of the ADHD-T-Dari was performed. If Cronbach’s alpha exceeded 0.750, it was deemed that the criteria for internal consistency were met. The main component analysis was used to establish the one-, two-, and three-dimensionality of the ADHD-T-Dari.

To assess concept validity, an exploratory factor analysis (EFA) with Varimax rotation was performed. It was determined whether the ADHD-T-Dari structure, which should correspond to the structure of the original ADHD-T version, was one with a single item or one with many items. Kaiser (own value) and Cattell (scree plot) were used as the two criteria for separating the number of items.

When deciding which items to include in each factor, it was decided from the outset that items that loaded more than 0.40 on a factor would be included. Ten respondents per item is the recommended minimum sample size.

The accuracy of the adaptation of the obtained results to the imposed structure of ADHD-T-Dari, resulting from the theoretical assumptions or another structure derived from the EFA, was assessed using confirmatory factor analysis (CFA). The proposed indices should have the following values: goodness of fit index (GFI), comparative fit index (CFI) and Tucker-Lewis index (TLI) > 0.90 [12]; root mean squared error of approximation (RMSEA) < 0.08 [13]; χ^2^ divided by degrees of freedom (χ^2^/*df* ratio) 3-5 [14]. Microsoft Excel and IBM SPSS 28 were used for the calculation of all statistical tests. IMB AMOS version 28 was used for the CFA calculations.

### Ethical considerations

Parents reviewed the parental consent form, obtained their responses regarding the study, and confirmed their consent. Baseline data was collected in individual interviews. Baseline data included the socio-demographic information provided by the parents when completing the parental consent form. The Ethics Committee of Kabul University (No. 25, 03/03/2020) granted the permission to conduct this study.

## Results

### Socio-demographic characteristics

A total of 358 public primary school students with a mean age of 7.7 years (5.67-9.83 years) (SD = 0.93), of whom 48.6% were male and 51.4% female, participated in the study. The families of the students have a low (57%) or medium income (43%). 75.7% of mothers and 56.4% of fathers were illiterate. 65.6% of the students were Tajik, followed by 43% Pashtun, 4.5% Sadat, and 2.5% Hazara, while the remaining students did not specify their ethnicity.

### Content and face validities

The results for the I-CVI of ADHD-T-Dari ranged from 0.875 to 1. The S-CVI of ADHD-T-Dari was at a satisfactory level. All 18 items of the ADHD-T-Dari were submitted for validation as none of them were removed or modified. The I-FVI of ADHD-T-Dari was 0.975 to 1, indicating an excellent comprehension for all 18 items. Minor corrections and fine-tuning of the questionnaire were made according to their comments and recommendations. The results of CVI and FVI for inattention and hyperactivity-impulsivity of ADHD-T-Dari are presented in S1, S2. S3 and S4 Tables.

### Construct validity

The analysis of the raw data revealed the validity of the predictions of the exploratory factor analysis (EFA). The correlation matrix’s determinant for the ADHD-T-Dari items was close to zero (0.001), indicating significant collinearity between the inattention and hyperactivity-impulsivity domains of the items. According to the Bartlett’s test of sphericity, the matrix with the coefficients of correlation was a unit matrix (χ^2^ = 2326.36, *df* = 153, *p* < 0.001). The Kaiser-Meyer-Olkin index, which gauges sampling adequacy, was 0.913, indicating that the parameter satisfied the assumptions (> 0.5). The 18-item EFA test of ADHD-T-Dari was divided into four components, which together accounted for 46.58% of the overall variance. Nevertheless, the scree plot of this result showed a two-factor solution that accounted for 46.58% of the total variance. The other factor had only 1.020 and 1.005 initial eigenvalues (5.968%), which were not included in the first component analysis (Fig 2). The two factors (Inattention, hyperactivity-impulsivity) loading of ADHD-T-Dari were higher than 0.4 (S5 Table).

**Fig 2.**
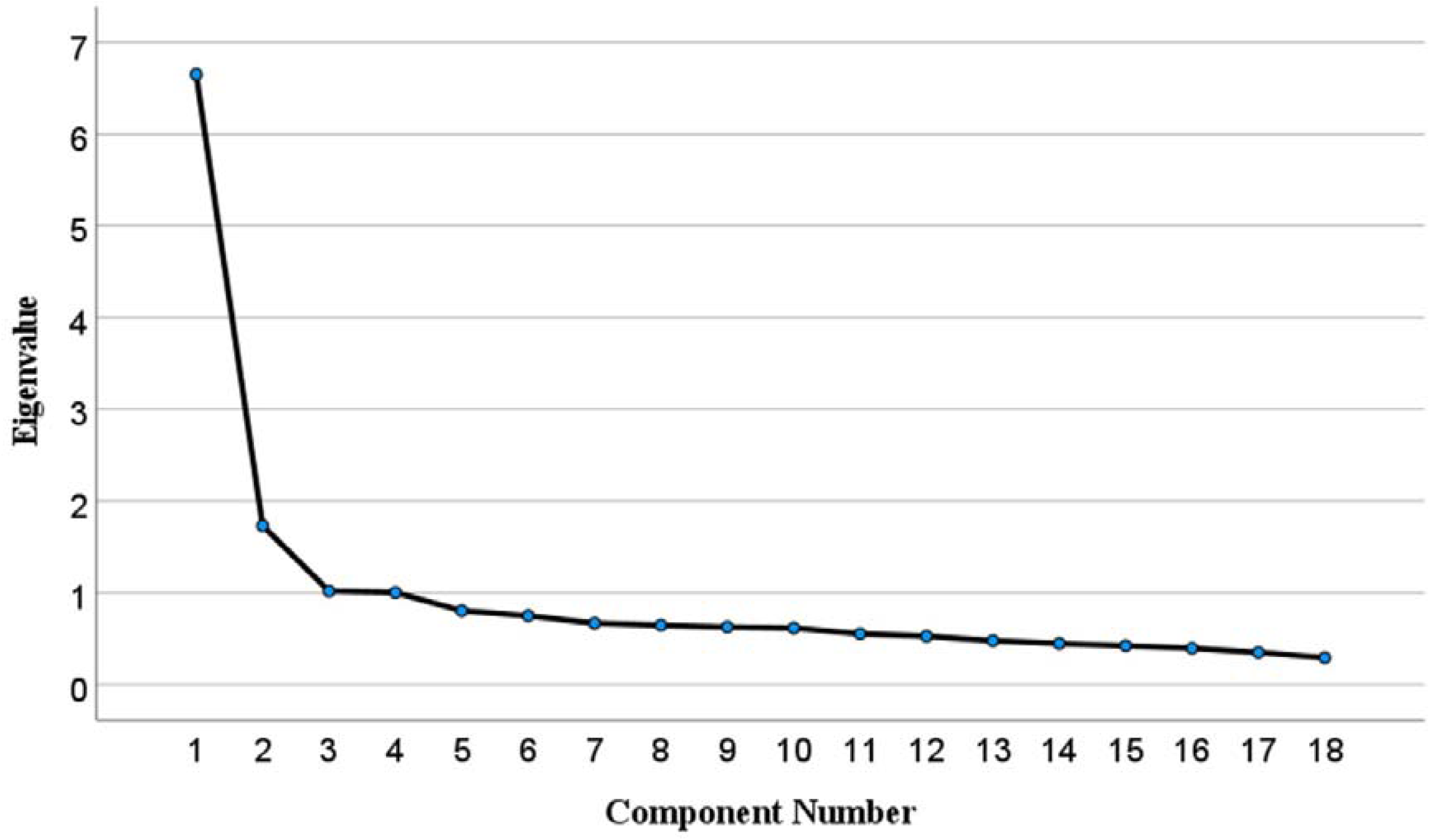
ADHD-T-Dari factor loading scree plot for Eigenvalue.

### Confirmatory factor analysis (CFA)

CFA was used to compare the one-dimensional goodness-of-fit and the two- and three-dimensional models of ADHD-T-Dari for the data collected. Compared to the two-dimensional version, the findings for the one- and three-dimensional versions of ADHD-T were less acceptable. The analysis revealed that the chi-square statistic on the degrees of freedom (χ^2^/*df*) of the two-dimensional model was 2.501 (χ^2^ = 335.147, *df* = 134). The RMSEA was 0.065. The CFI score was 0.909 and the GFI was 0.905. The CFI and GFI values indicate a good fit of the model. As a result, we consider our two-factor model to be accepted. Table 1 lists the fit indices for all CFA models, and Figs 3,4 and 5 show the standardised factor loadings for the one-, two- and three-factor models of ADHD-T-Dari.

**Fig 3.**
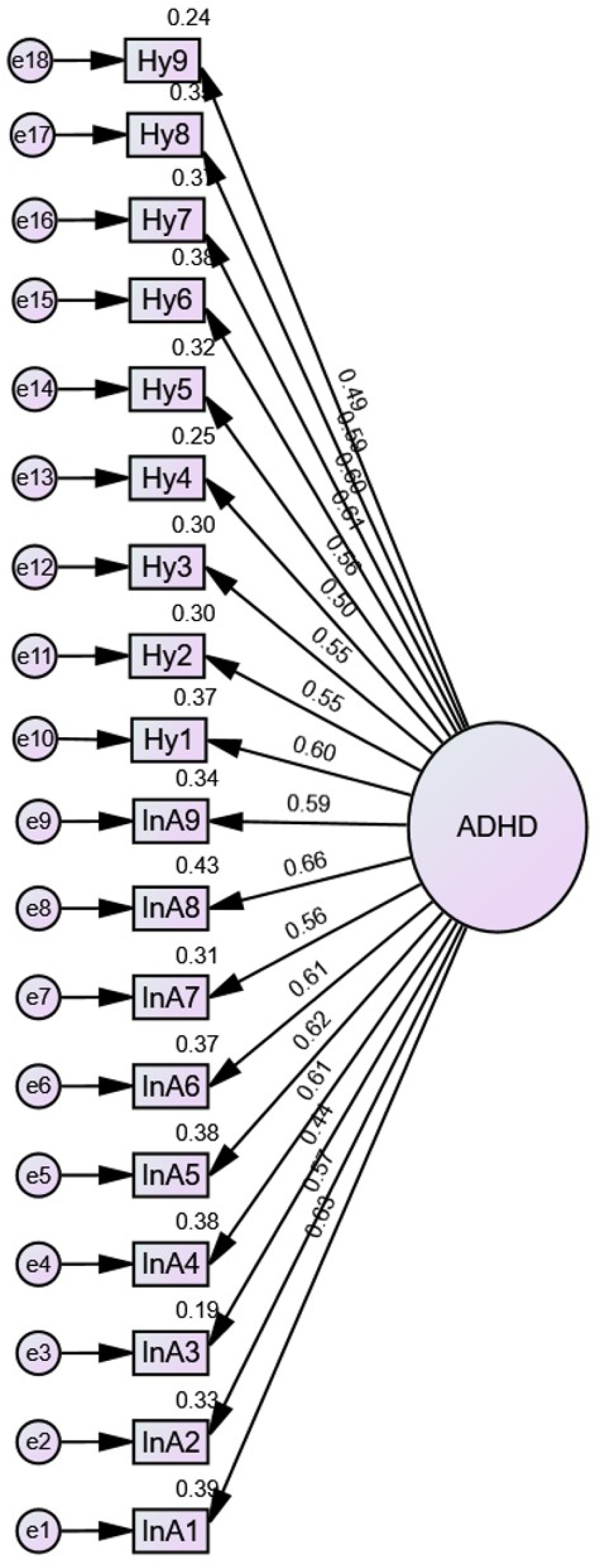
The CFA of the ADHD-T-Dari single-factor model with its loading. The one-factor model is depicted in a CFA diagram (ADHD-T-5.es dimensions). Each rectangle (Hy = hyperactivity-impulsivity, InA = inattention) and oval circle (ADHD) denote a single item or dimension, respectively. Each circle is associated with items that define only one dimension, not with the items that define the other dimension (average item-subscale correlation). Arrows are used to depict correlations between dimensions and items.

**Fig 4.**
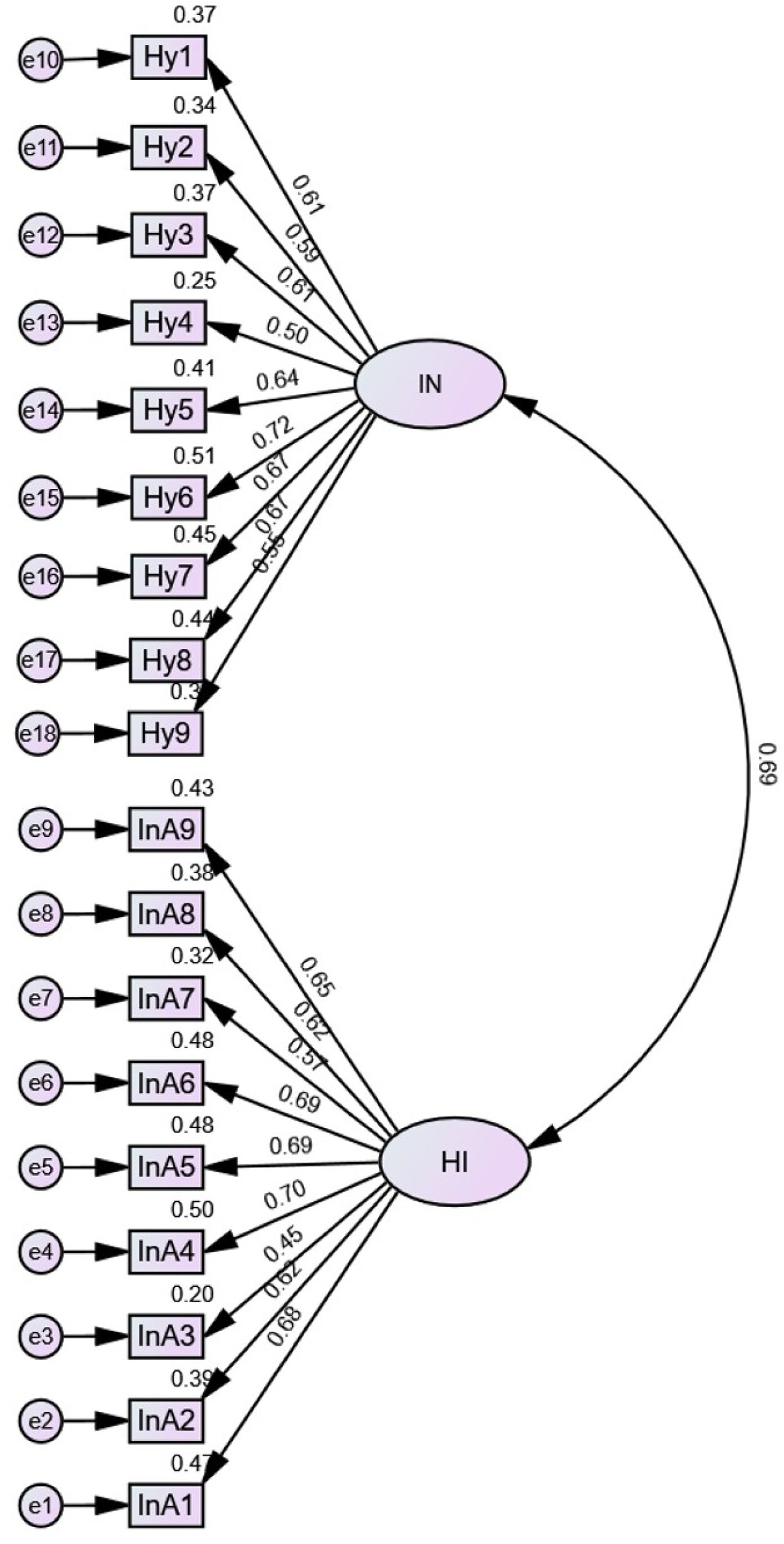
The CFA of the ADHD-T-Dari two-factors model with its loading. The two-factor model and the correlations between the two subscales are depicted in a CFA diagram (ADHD-T-Dari dimensions). Each rectangle (Hy = hyperactivity-impulsivity, InA = inattention) and each oval circle (IN = inattention, HI = hyperactivity-impulsivity) denote a single item and dimension. Each circle is associated with the items that define only one dimension, not with the items that define the other dimension (average item-subscale correlation). Double arrows are used to depict correlations between dimensions and/or items (correlation coefficient between item/inter-subscales).

**Fig 5.**
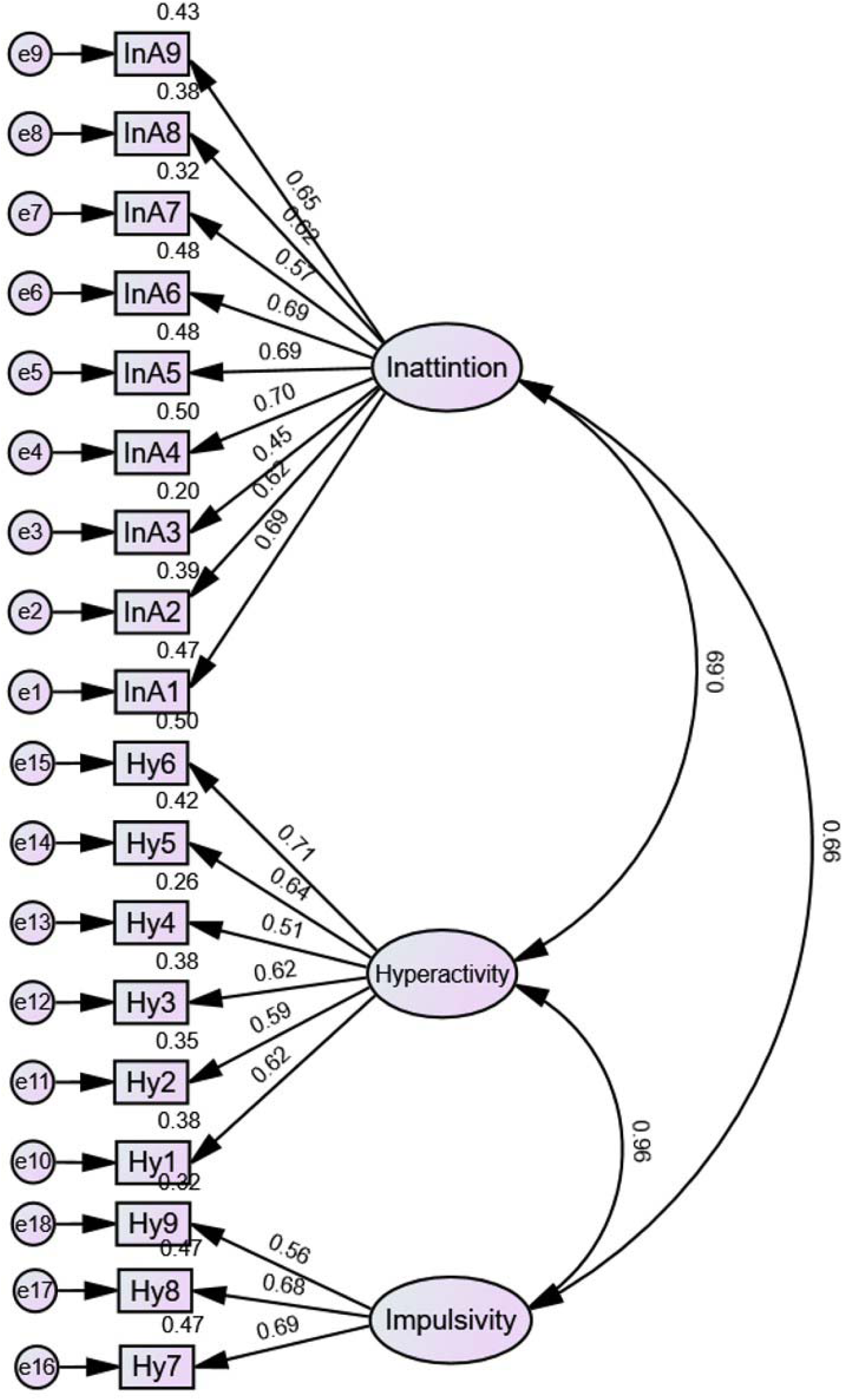
The CFA of the ADHD-T-Dari three-factors model with its loading. The three-factor model and the correlations between the three subscales are depicted in a CFA diagram (ADHD-T-5.es dimensions). Each rectangle (Hy = hyperactivity-impulsivity, InA = inattention) and each oval circle (inattention, hyperactivity-impulsivity) denote a single item and dimension, respectively. Each circle is linked to the items that define only one dimension, not to the items that define the other dimension (average item-subscale correlation). Double arrows are used to depict correlations between dimensions and/or items (correlation coefficient between items/inter-subscales).

**Table 1.** The result of CFA for one-, two- and three factors of ADHD-T-Dari.

| Fit indices | *p* | CMIN/*df* | GFI | CFI | TLI | RMSEA |
| --- | --- | --- | --- | --- | --- | --- |
| **Recommended** | Insignificant | 3-5 | >0.90 | >0.90 | >0.90 | <0.08 |
| **One factor** | <0.001 | 4.25 | 0.809 | 0.802 | 0.776 | 0.095 |
| **Two factors** | <0.001 | 2.501 | 0.905 | 0.909 | 0.896 | 0.065 |
| **Three factors** | <0.001 | 2.525 | 0.907 | 0.909 | 0.895 | 0.065 |

Note. CMIN = Chi-Square, *df* = degree of freedom, GFI = goodness of the fit index, CFI = comparative fit index, TLI = Tucker-Lewis's index, RMSEA = root mean square error of approximation.

### Reliability

The overall Cronbach’s alpha for the 18 items of the ADHD-T-Dari was 0.898, while the components measuring inattention and hyperactivity-impulsivity were 0.851 and 0.847, respectively. This indicates that the items form a scale with a reasonable degree of internal consistency and reliability and are higher than the predicted 0.75 limits. In this study group, the average score for the 18 ADHD-T-Dari items was 8.01 (*SD* = 8.47). These 18 items have an adjusted item-total correlation of over 0.40, which is considered acceptable (Table 2).

**Table 2.** Cronbach’s alpha and descriptive statistics for 18 ADHD-T-Dari if items deleted.

| **Items** | **Mean** | ***SD*** | **Item-Total**<br>**Correlation** | **Cronbach's Alpha**<br>**if Item Deleted** |
| --- | --- | --- | --- | --- |
| **InA1** | 0.42 | 0.642 | 0.578 | 0.892 |
| **InA2** | 0.35 | 0.684 | 0.527 | 0.893 |
| **InA3** | 0.32 | 0.682 | 0.405 | 0.896 |
| **InA4** | 0.30 | 0.615 | 0.566 | 0.892 |
| **InA5** | 0.38 | 0.707 | 0.564 | 0.892 |
| **InA6** | 0.42 | 0.731 | 0.558 | 0.892 |
| **InA7** | 0.55 | 0.854 | 0.519 | 0.893 |
| **InA8** | 0.48 | 0.826 | 0.619 | 0.890 |
| **InA9** | 0.43 | 0.770 | 0.536 | 0.892 |
| **Hy1** | 0.57 | 0.898 | 0.581 | 0.891 |
| **Hy2** | 0.43 | 0.778 | 0.536 | 0.892 |
| **Hy3** | 0.38 | 0.760 | 0.531 | 0.893 |
| **Hy4** | 0.30 | 0.709 | 0.479 | 0.894 |
| **Hy5** | 0.46 | 0.831 | 0.546 | 0.892 |
| **Hy6** | 0.73 | 0.952 | 0.596 | 0.891 |
| **Hy7** | 0.70 | 0.939 | 0.583 | 0.891 |
| **Hy8** | 0.44 | 0.803 | 0.575 | 0.891 |
| **Hy9** | 0.35 | 0.728 | 0.477 | 0.894 |

Note. InA= inattention, Hy= hyperactivity-impulsivity

### Age and gender difference of ADHD

The findings of the independent *t*-test using the ADHD-T-Dari scores as the dependent variables and the child’s gender as the independent variable, are summarised in Table 3. The great impacts of gender were found to be statistically significant, with males being categorised as more inattentive, hyperactive and impulsive than girls. The effect sizes were often very large. Table 4 presents the results of the one-way ANOVA for age and respondents, for the inattention, hyperactivity-impulsivity and overall ADHD scales. In general, the effect sizes were small.

**Table 3.** ADHD-T-Dari descriptive and independent tests for 174 male and 184 female students.

| Variables | Male |  | Female |  | *t* (356) | *p* | Cohen's *d* |
| --- | --- | --- | --- | --- | --- | --- | --- |
|  | *M* | *SD* | *M* | *SD* |  |  |  |
| **Inattention** | 10.23 | 6.69 | 5.12 | 5.84 | 7.713 | <0.001 | 0.81 |
| **Hyperactivity** | 10.30 | 4.32 | 4.29 | 5.87 | 8.585 | <0.001 | 1.16 |
| **ADHD** | 20.53 | 12.42 | 9.41 | 10.15 | 9.297 | <0.001 | 0.98 |

**Table 4.**
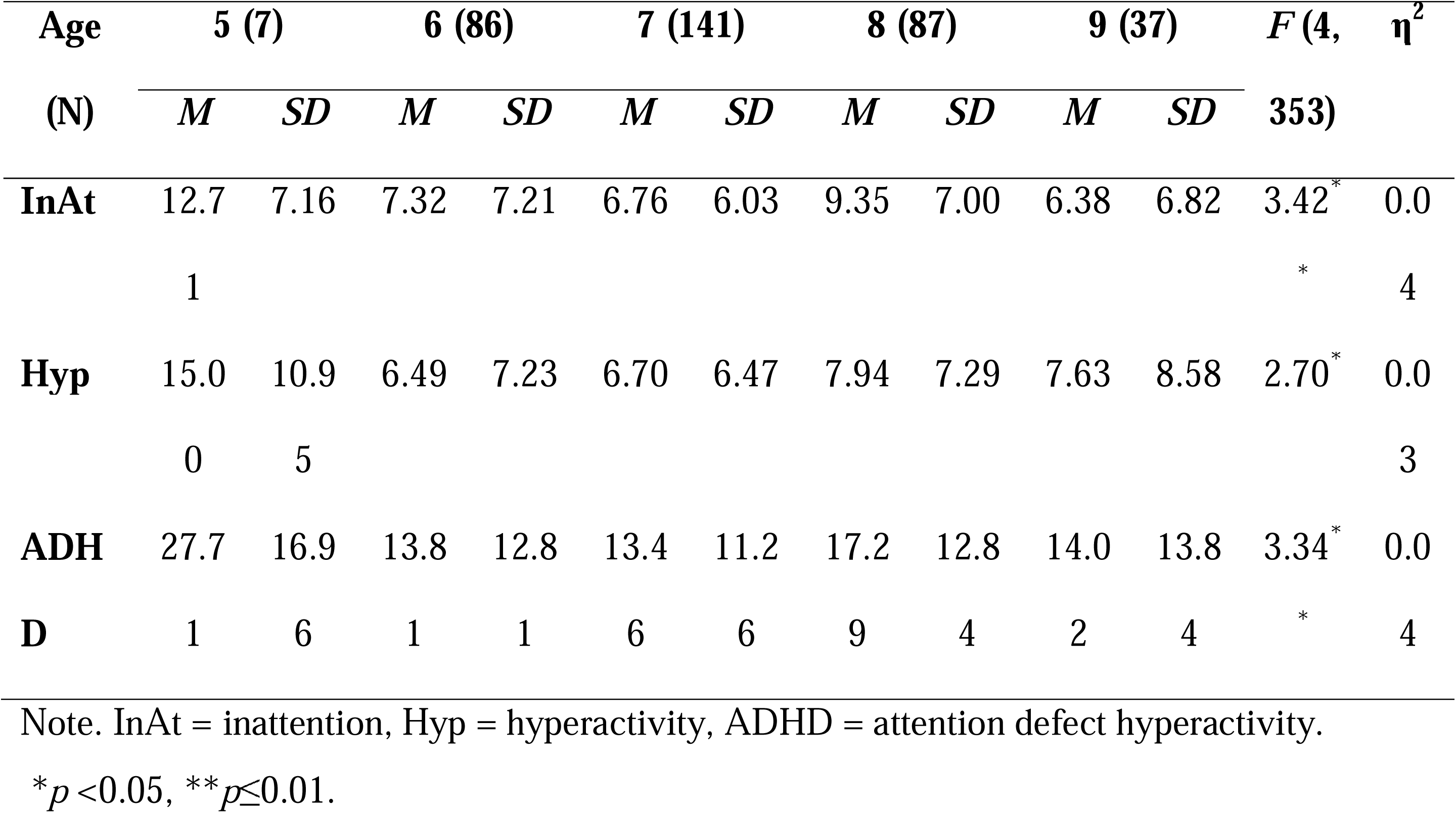
One-Way ANOVA of ADHD for age groups of students.

| Age<br>(N) | 5 (7) | | 6 (86) | | 7 (141) | | 8 (87) | | 9 (37) | | *F* (4,<br>353) | $\eta^2$ |
| --- | --- | --- | --- | --- | --- | --- | --- | --- | --- | --- | --- | --- |
|  | *M* | *SD* | *M* | *SD* | *M* | *SD* | *M* | *SD* | *M* | *SD* |  |  |
| **InAt** | 12.7 | 7.16 | 7.32 | 7.21 | 6.76 | 6.03 | 9.35 | 7.00 | 6.38 | 6.82 | 3.42* | 0.0 |
|  | 1 |  |  |  |  |  |  |  |  |  | * | 4 |
| **Hyp** | 15.0 | 10.9 | 6.49 | 7.23 | 6.70 | 6.47 | 7.94 | 7.29 | 7.63 | 8.58 | 2.70* | 0.0 |
|  | 0 | 5 |  |  |  |  |  |  |  |  |  | 3 |
| **ADHD** | 27.7 | 16.9 | 13.8 | 12.8 | 13.4 | 11.2 | 17.2 | 12.8 | 14.0 | 13.8 | 3.34* | 0.0 |
| **D** | 1 | 6 | 1 | 1 | 6 | 6 | 9 | 4 | 2 | 4 | * | 4 |

Note. InAt = inattention, Hyp = hyperactivity, ADHD = attention defect hyperactivity.
\* $p < 0.05$ , \*\* $p \leq 0.01$ .

## Discussion

This study aimed to describe the prevalence of ADHD in public primary school students in Kabul City and to validate the Dari-language version of ADHD-T. However, to the best of our knowledge, this is the first study conducted in Afghanistan that adapts a questionnaire for teachers to utilise in determining whether their students have ADHD. The finalised validation indicates strong psychometric characteristics of the ADHD-T-Dari, including validity and reliability. However, in line with previous validation studies, no variations from the original ADHD-T version were found. All items were validated, according to the judgement of the expert panels.

The standard goodness-of-fit indices for ADHD demonstrated that the bi-factor models provided better agreement, which is consistent with some studies [7]. However, several studies on earlier forms of ADHD have shown a tendency towards high goodness-of-fit indices for single-factor models [15]. The two and three ADHD components were evident in the current study when one-, two-, and three-factorial analyses were used. The two-factor was significant in the general population sample confirming previous findings [8]. The original version of ADHD-T and ADHD-T-Dari were identical. The Kaiser criterion indicated a two-item structure of the scale when examining the factor structure of the ADHD-T-Dari’s, and other research also claimed a two-component solution [3,8]. A two-factor structure of the ADHD-T-Dari was found to be more effective for the CFA than a single- and a three-factor structure.

The TLI for two-factor models in our study was somewhat (0.986) less than the suggested value of > 0.90 but was still higher than for one-(0.776) and three-factor (0.895) models. However, there are no “golden rules” for the TLI. When utilising the practical fit indices to evaluate model fit in empirical studies, researchers should consider the number of observed variables [16]. A relative decrease in misfit is measured by the TLI [17] per degree of freedom. When the model was mis specified by fitting a single-factor model to two-factor data or by removing cross-loadings, the population RMSEA tended to decrease as p increased, and the population values for CFI and TLI tended to decrease. However, it was found that the population values of CFI and TLI tended to increase when *p* increased [18].

Our findings indicate that the scale is sufficiently reliable (e.g. internal consistency). The Cronbach’s alpha for the inattention domain with 9 items of ADHD-T-Dari was = 0.851, for hyperactivity-impulsivity with 9 items was 0.847 and for a total of 18 items was 0.898, which was above the recommended value of 0.75 and showed a strong internal consistency. These results are consistent with the source of the questionnaire [7] and the findings of multinational studies [8,19] that have examined the validity of the teacher version of ADHD-5. The findings of the psychometric investigation of the ADHD-T-Dari suggest that the scale can be used by teachers in practice as an instrument for evaluating ADHD and academic performance in school-aged children.

On average, ADHD affects between 2% and 7% of the world’s population [1,2], and in a meta-analysis, the pooled prevalence was reported to be between 1.1 and 16.7% in Italy only [20]. In this study, the prevalence of ADHD was 21.5%, well above the global average. However, there are no previous studies demonstrating the prevalence of ADHD in the Afghan population. This increased prevalence of ADHD could be related to the younger age of the study population and urbanisation. According to a meta-analysis, ADHD is more common in both sexes up to the age of 9 years and decreases thereafter [21]. In terms of habitation, ADHD is more common in urban areas [22]. According to studies, males are more likely to have ADHD than females [23]. The current study confirms this trend, with boys being much more likely to have ADHD than girls (23.6 vs. 19.6%).

This study has several limitations. Firstly, only teachers answered the questionnaire. Our findings cannot be extrapolated to what the scale would say if it had been completed by parents. Second, we only included a sample of students between the ages of 6 and 9, as the original questionnaire was designed for children between the ages of 6 and 17. Third, due to the larger number of classes and the teachers’ lack of experience with psychometric measurement methods in the present study, it was difficult for a single teacher to focus on one student and complete the scale, even though the factor loadings are lower and the residual variances are higher.

## Conclusion

The results of the psychometric examination of the ADHD-T-Dari show that the validity and reliability of the instruments are of high quality and can be used for the assessment of ADHD in school-age children.

## Data Availability

All data produced in the present study work are contained in the manuscript

## Acknowledgements

We would like to thank Dr. George J. DuPaul for giving permission to use the ADHD rating Scale-5. We would like to acknowledge the academic staff from the Department of Counselling and the Department of Psychology at the Faculty of Psychological Science, Kabul University, for their assistance with content validity. We would also like to express our gratitude to the Department of English Language at Kabul University, as well as Mariam Momand, Bushra Wali, and Sheela Shneezai for their help in forward and backward translation. Our appreciation goes to the teachers of Bibi Sara High School who assisted with face validity and the students (Bibi Marwa Hashimi, Maryam Zarifi, Aryan Muqadas Noorzad, Sahar Mhalligy, Umefarwa Haidari, Nastaran Haidari, Parivash Khanzadah, Shila Joyan, Arshia Moddaa, Sadam Sherzai, Yad Mohammad Nazary, Sediqa Mirzad and Hela Yaqub) from the Faculty of Pharmacy, Kabul University, who assisted in data collection. We would like to thank all teachers, students, and parents who participated in the study and gave their consent.

## Supporting information

**S1 Table.**
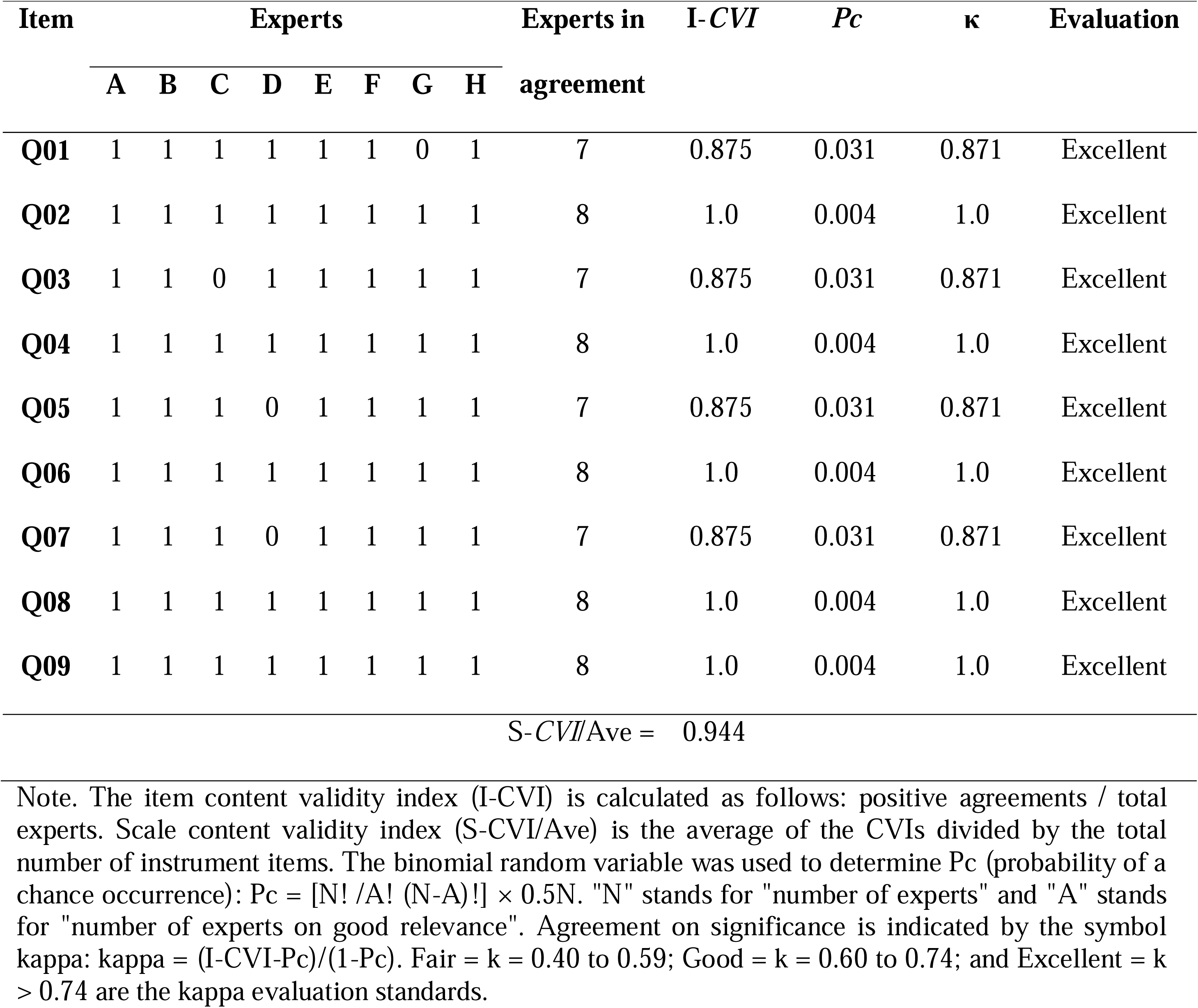
The scores of 8 experts on 9 items of the Inattention domain of ADHD-T-Dari: CVI.

**S2 Table.**
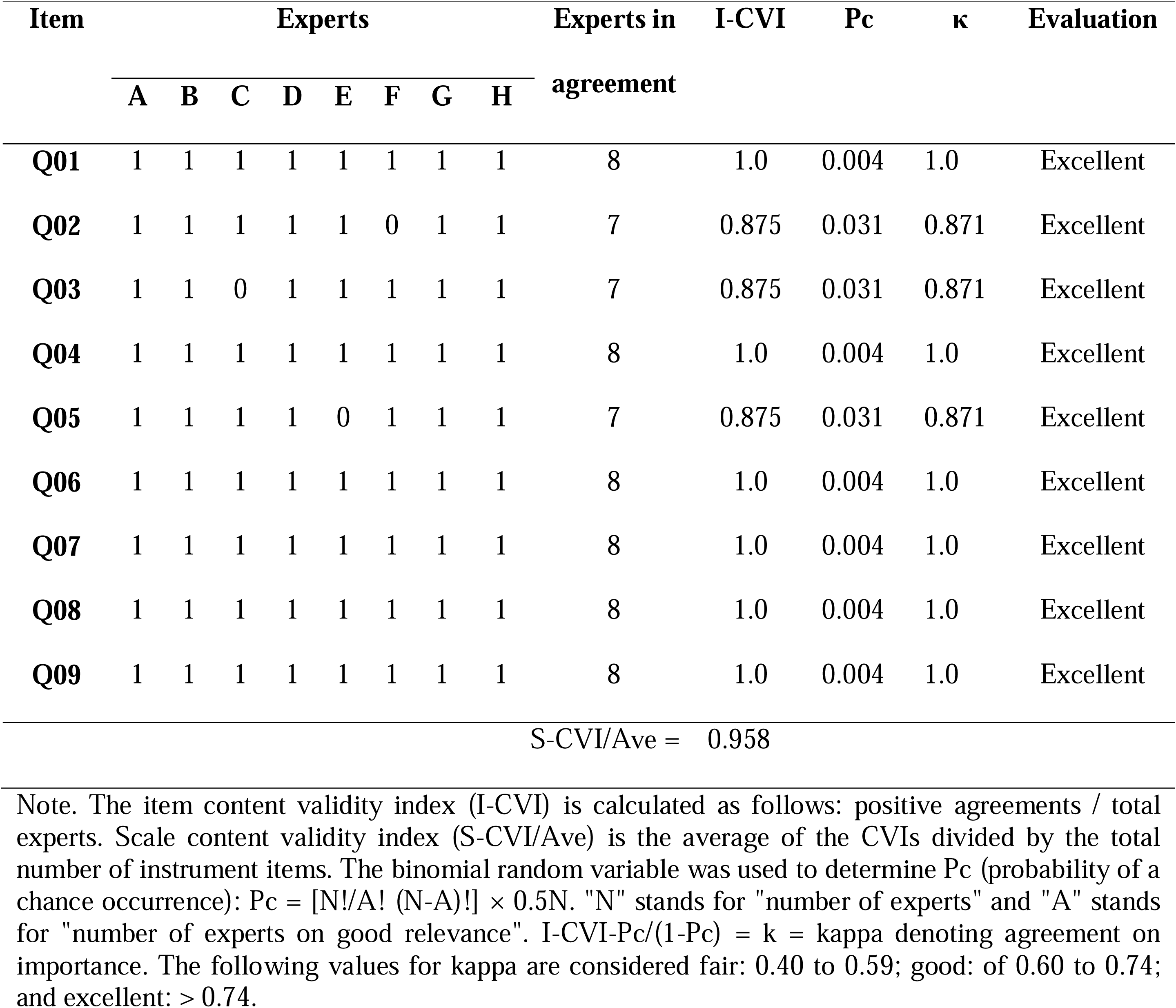
The scores of 8 experts on 9 items of hyperactivity/Impulsivity domain of ADHD-T-Dari: CVI.

**S3 Table.**
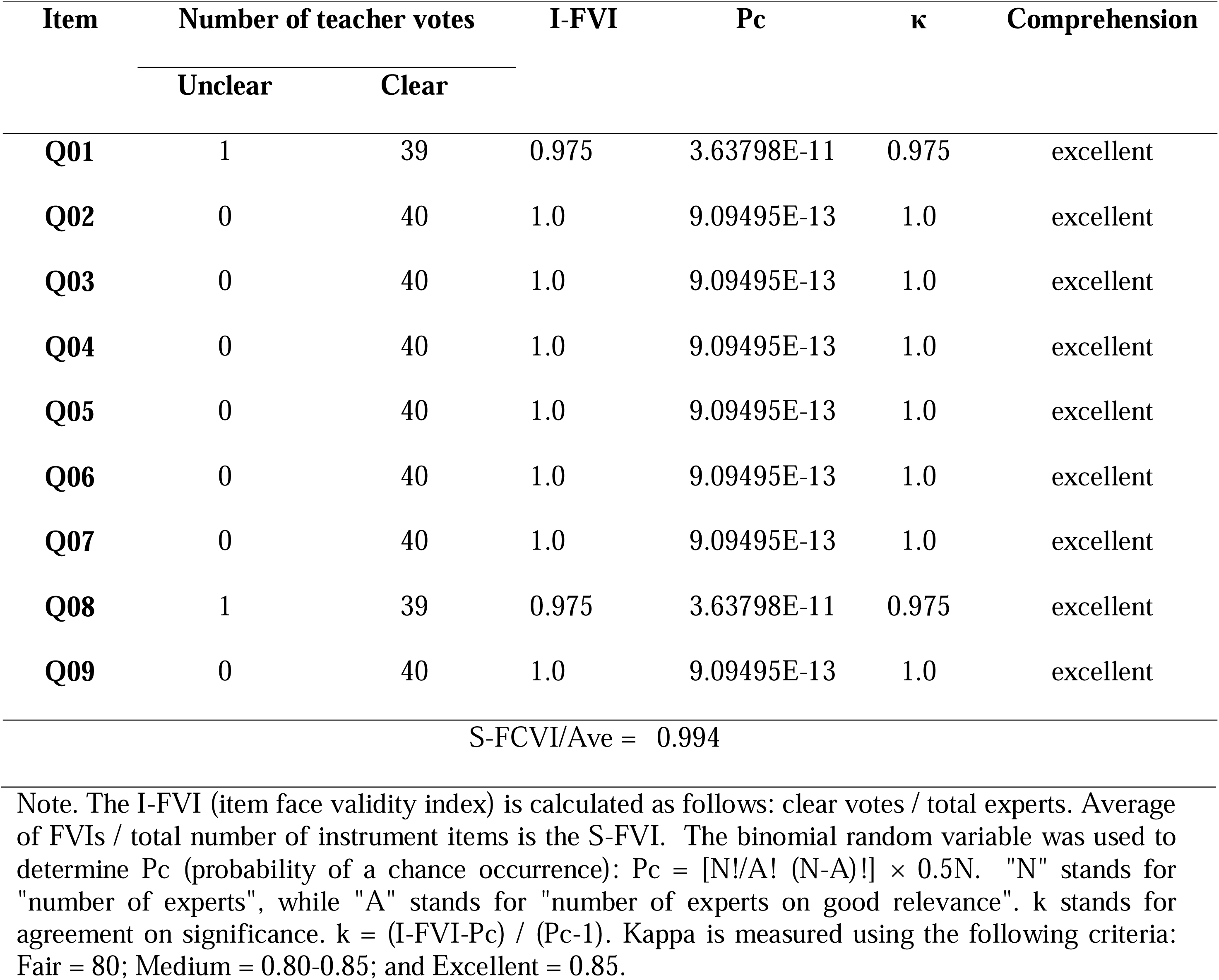
The scores of 40 teachers on 9 items of the Inattention domain of ADHD-T-Dari: FVI.

**S4 Table.**
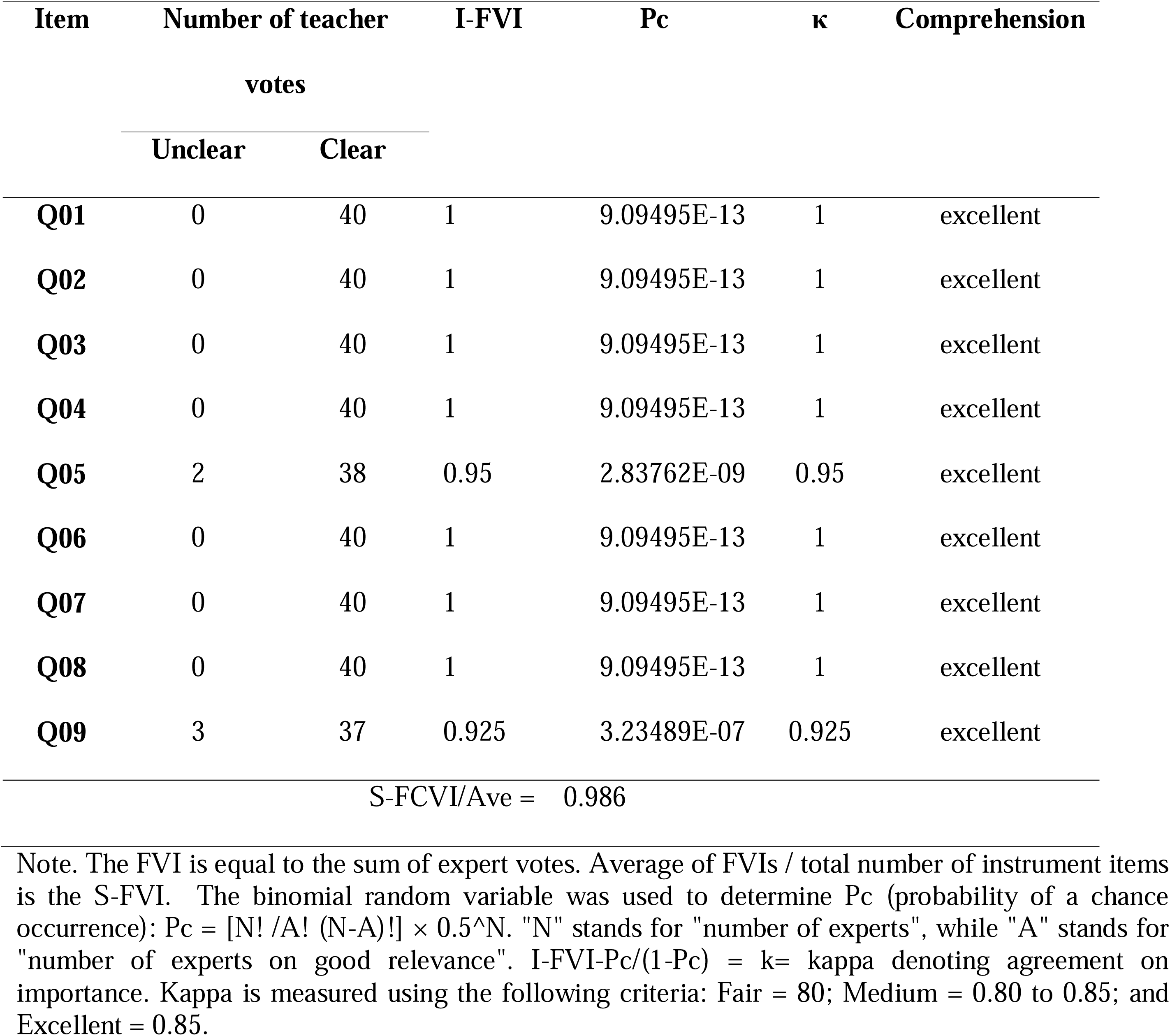
The scores of 40 teachers on 9 items of hyperactivity/Impulsivity domain of ADHD-T-Dari: FVI.

**S5 Table.**
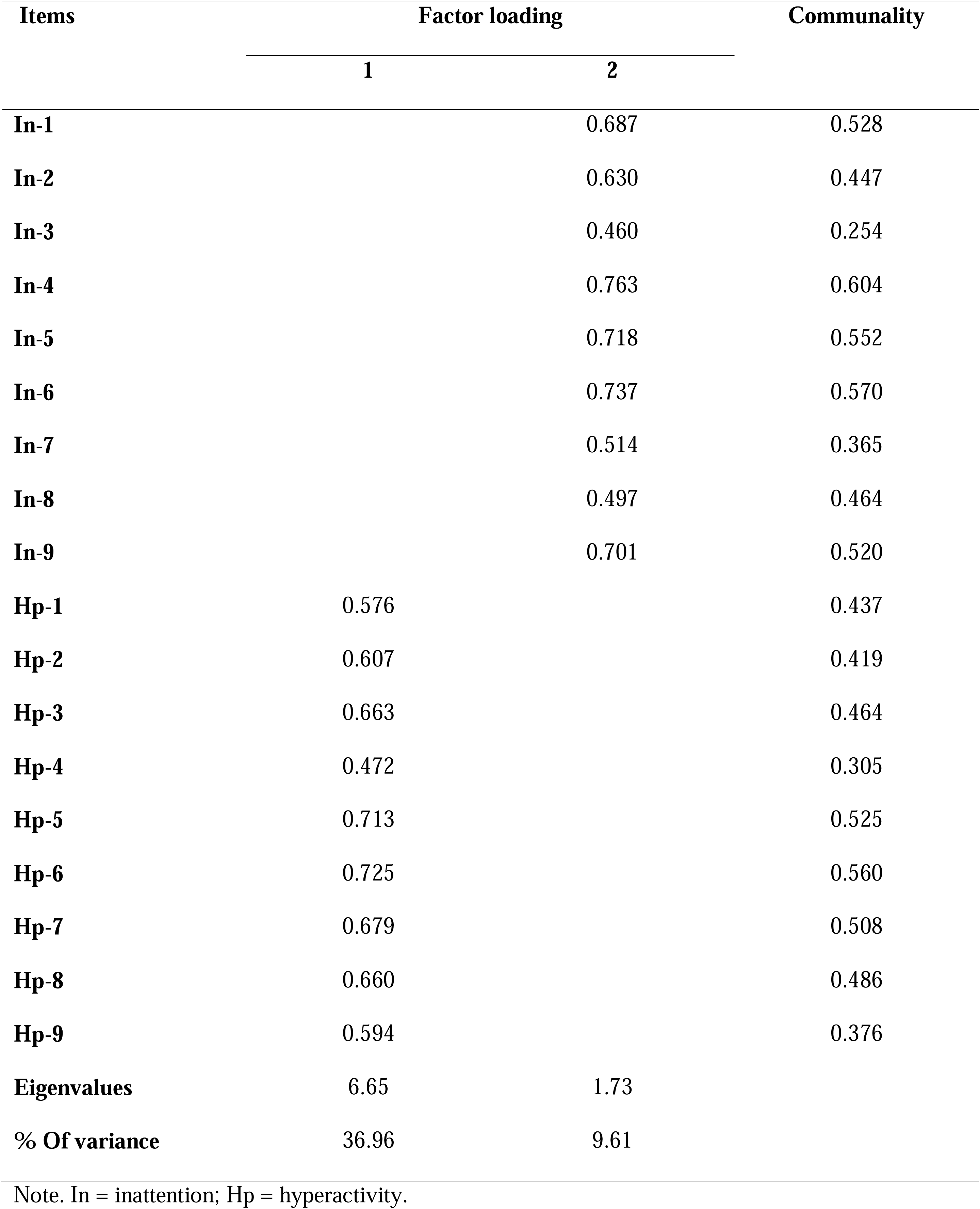
Principal Component Factor Analysis for ADHD using Varimax Rotation Factor Loadings.

