## Supplementary 1 for "Psychometric characteristics of ADHD-T-5 in its Dari version among public primary school students in Kabul City, Afghanistan"

**S1 Table. The scores of 8 experts on 9 items of Inattention domain of ADHD-T-Dari: CVI.**

| Item | Experts | | | | | | | | Experts in agreement | I-*CVI* | *Pc* | κ | Evaluation |
|---|---|---|---|---|---|---|---|---|---|---|---|---|---|
| | A | B | C | D | E | F | G | H | | | | | |
| **Q01** | 1 | 1 | 1 | 1 | 1 | 1 | 0 | 1 | 7 | 0.875 | 0.031 | 0.871 | Excellent |
| **Q02** | 1 | 1 | 1 | 1 | 1 | 1 | 1 | 1 | 8 | 1.0 | 0.004 | 1.0 | Excellent |
| **Q03** | 1 | 1 | 0 | 1 | 1 | 1 | 1 | 1 | 7 | 0.875 | 0.031 | 0.871 | Excellent |
| **Q04** | 1 | 1 | 1 | 1 | 1 | 1 | 1 | 1 | 8 | 1.0 | 0.004 | 1.0 | Excellent |
| **Q05** | 1 | 1 | 1 | 0 | 1 | 1 | 1 | 1 | 7 | 0.875 | 0.031 | 0.871 | Excellent |
| **Q06** | 1 | 1 | 1 | 1 | 1 | 1 | 1 | 1 | 8 | 1.0 | 0.004 | 1.0 | Excellent |
| **Q07** | 1 | 1 | 1 | 0 | 1 | 1 | 1 | 1 | 7 | 0.875 | 0.031 | 0.871 | Excellent |
| **Q08** | 1 | 1 | 1 | 1 | 1 | 1 | 1 | 1 | 8 | 1.0 | 0.004 | 1.0 | Excellent |
| **Q09** | 1 | 1 | 1 | 1 | 1 | 1 | 1 | 1 | 8 | 1.0 | 0.004 | 1.0 | Excellent |
| | | | | | | | | | S-*CVI*/Ave = | 0.944 | | | |

Note. The item content validity index (I-CVI) is calculated as follows: positive agreements / total experts. Scale content validity index (S-CVI/Ave) is the average of the CVIs divided by the total number of instrument items. The binomial random variable was used to determine Pc (probability of a chance occurrence): Pc = [N! /A! (N-A)!] × 0.5N. "N" stands for "number of experts" and "A" stands for "number of experts on good relevance". Agreement on significance is indicated by the symbol kappa: kappa = (I-CVI-Pc)/(1-Pc). Fair = k = 0.40 to 0.59; Good = k = 0.60 to 0.74; and Excellent = k > 0.74 are the kappa evaluation standards.
