## Supplementary 3 for "Psychometric characteristics of ADHD-T-5 in its Dari version among public primary school students in Kabul City, Afghanistan"

**S3 Table. The scores of 40 teachers on 9 items of Inattention domain of ADHD-T-Dari: FVI.**

| Item | Number of teacher votes | | I-FVI | Pc | κ | Comprehension |
|---|---|---|---|---|---|---|
| | Unclear | Clear | | | | |
| **Q01** | 1 | 39 | 0.975 | 3.63798E-11 | 0.975 | excellent |
| **Q02** | 0 | 40 | 1.0 | 9.09495E-13 | 1.0 | excellent |
| **Q03** | 0 | 40 | 1.0 | 9.09495E-13 | 1.0 | excellent |
| **Q04** | 0 | 40 | 1.0 | 9.09495E-13 | 1.0 | excellent |
| **Q05** | 0 | 40 | 1.0 | 9.09495E-13 | 1.0 | excellent |
| **Q06** | 0 | 40 | 1.0 | 9.09495E-13 | 1.0 | excellent |
| **Q07** | 0 | 40 | 1.0 | 9.09495E-13 | 1.0 | excellent |
| **Q08** | 1 | 39 | 0.975 | 3.63798E-11 | 0.975 | excellent |
| **Q09** | 0 | 40 | 1.0 | 9.09495E-13 | 1.0 | excellent |
| S-FCVI/Ave = 0.994 | | | | | | |

Note. The I-FVI (item face validity index) is calculated as follows: clear votes / total experts. Average of FVIs / total number of instrument items is the S-FVI. The binomial random variable was used to determine Pc (probability of a chance occurrence): Pc = [N!/A! (N-A)!] × 0.5N. "N" stands for "number of experts", while "A" stands for "number of experts on good relevance". k stands for agreement on significance. k = (I-FVI-Pc) / (Pc-1). Kappa is measured using the following criteria: Fair = 80; Medium = 0.80-0.85; and Excellent = 0.85.
