## Supplementary 4 for "Psychometric characteristics of ADHD-T-5 in its Dari version among public primary school students in Kabul City, Afghanistan"

**S4 Table. The scores of 40 teachers on 9 items of hyperactivity/Impulsivity domain of ADHD-T-Dari: FVI.**

| Item | Number of teacher votes | | I-FVI | Pc | κ | Comprehension |
|---|---|---|---|---|---|---|
| | Unclear | Clear | | | | |
| **Q01** | 0 | 40 | 1 | 9.09495E-13 | 1 | excellent |
| **Q02** | 0 | 40 | 1 | 9.09495E-13 | 1 | excellent |
| **Q03** | 0 | 40 | 1 | 9.09495E-13 | 1 | excellent |
| **Q04** | 0 | 40 | 1 | 9.09495E-13 | 1 | excellent |
| **Q05** | 2 | 38 | 0.95 | 2.83762E-09 | 0.95 | excellent |
| **Q06** | 0 | 40 | 1 | 9.09495E-13 | 1 | excellent |
| **Q07** | 0 | 40 | 1 | 9.09495E-13 | 1 | excellent |
| **Q08** | 0 | 40 | 1 | 9.09495E-13 | 1 | excellent |
| **Q09** | 3 | 37 | 0.925 | 3.23489E-07 | 0.925 | excellent |
| | | | S-FCVI/Ave = | 0.986 | | |

Note. The FVI is equal to the sum of expert votes. Average of FVIs / total number of instrument items is the S-FVI. The binomial random variable was used to determine Pc (probability of a chance occurrence): Pc = [N! /A! (N-A)!] × 0.5^N. "N" stands for "number of experts", while "A" stands for "number of experts on good relevance". I-FVI-Pc/(1-Pc) = k= kappa denoting agreement on importance. Kappa is measured using the following criteria: Fair = 80; Medium = 0.80 to 0.85; and Excellent = 0.85.
