## Supplementary 5 for "Psychometric characteristics of ADHD-T-5 in its Dari version among public primary school students in Kabul City, Afghanistan"

**S5 Table. Principal Component Factor Analysis for ADHD using Varimax Rotation Factor Loadings.**

| Items | Factor loading | | Communality |
|---|---|---|---|
| | 1 | 2 | |
| **In-1** | | 0.687 | 0.528 |
| **In-2** | | 0.630 | 0.447 |
| **In-3** | | 0.460 | 0.254 |
| **In-4** | | 0.763 | 0.604 |
| **In-5** | | 0.718 | 0.552 |
| **In-6** | | 0.737 | 0.570 |
| **In-7** | | 0.514 | 0.365 |
| **In-8** | | 0.497 | 0.464 |
| **In-9** | | 0.701 | 0.520 |
| **Hp-1** | 0.576 | | 0.437 |
| **Hp-2** | 0.607 | | 0.419 |
| **Hp-3** | 0.663 | | 0.464 |
| **Hp-4** | 0.472 | | 0.305 |
| **Hp-5** | 0.713 | | 0.525 |
| **Hp-6** | 0.725 | | 0.560 |
| **Hp-7** | 0.679 | | 0.508 |
| **Hp-8** | 0.660 | | 0.486 |
| **Hp-9** | 0.594 | | 0.376 |
| **Eigenvalues** | 6.65 | 1.73 | |
| **% Of variance** | 36.96 | 9.61 | |

Note. In = inattention; Hp = hyperactivity.
